# A note on COVID-19 seroprevalence studies: a meta-analysis using hierarchical modelling

**DOI:** 10.1101/2020.05.03.20089201

**Authors:** Jérôme Levesque, David W. Maybury

## Abstract

In recent weeks, several seroprevalence studies have appeared which attempt to determine the prevalence of antibodies against SARS-CoV-2 in the population of certain European and American locations. Many of these studies find an antibody prevalence comparable to the false positive rate of their respective serology tests and the relatively low statistical power associated with each study has invited criticism. To determine the strength of the signal, we perform a meta-analysis on the publicly available seroprevalence data based on Bayesian hierarchical modelling with Markov Chain Monte Carlo and Generalized Linear Mixed Modelling with prediction sampling. We examine studies with results from Santa Clara County (CA), Los Angeles County (CA), San Miguel County (CO), Chelsea (MA), the Comté de Genève (Switzerland), and Gangelt (Germany). Our results are in broad agreement with the conclusions of the studies; we find that there is evidence for non-trivial levels of antibody prevalence across all study locations. However, we also find that a significant probability mass exists for antibody prevalence at levels lower than the reported figures. The results of our meta-analysis on the recent seroprevalence studies point to an important and strongly suggestive signal.

## 1 Introduction

The latest weapon against COVID-19 is serology testing—a search for antibodies against SARS-CoV-2 in the population’s bloodstream. A high prevalence of antibodies in the population helps us better understand the likelihood of asymptomatic infections or infections with mild symptoms. Moreover, high antibody prevalence indicates that a significant fraction of the population has already been exposed, lowering estimates of the infection fatality rate, and providing possible clues about herd immunity. Given the importance of determining the societal-wide exposure to SARS-CoV-2, critically understanding the results from the seroprevalence studies represents a pressing concern.

The recent serology study [1] from Santa Clara County, in which the authors report that the Santa Clara area has an antibody prevalence of 1.5% (exact binomial 95CI 1.11-1.97%), has received sharp criticism [2, 3]. In particular, the combination of the small sample size, small effect size, and a competitive false positive rate led to a study with relatively low statistical power. But, given the potential importance of serology analysis to our understanding of COVID-19, we must extract all the available information that the data contain. We simply cannot afford to dismiss weak signals.

Along with the Santa Clara study, a number of other studies have recently appeared using different types of tests [4, 5, 6, 7, 8]. Results are disparate. For example, the study in Chelsea, Massachusetts suggests an antibody prevalence as high as 30%, in marked contrast to the low levels detected by the Santa Clara study. Given the early days of this type of research, and the unevenness of the infection rates across regions, a noisy picture does not surprise us.

In this note we perform a meta-analysis based on the data from seven different studies. We apply two methods: 1) Bayesian hierarchical modelling with Markov Chain Monte Carlo, 2) Generalized Linear Mixed Modelling (GLMM) with prediction sampling. Both methods lead to similar results. We find that there is evidence for non-trivial levels of antibodies in the populations across all the studies—in broad concordance with the seroprevalence study conclusions—but that smaller than publicly stated levels are also probable.

## 2 Data, methods, and results

The studies we analyze were conducted in the last few weeks; peer-reviewed scientific publications are not yet available. We construct our datasets from scientific preprints, manufacturers’ specifications, and transcripts from interviews with study authors published in the news media.

Each study differs in its sampling strategy, and data on sample demographics is not yet available for most studies. In each study, the authors aim to collect a representative sample of their population. We did not attempt to post-stratify any of the results, and our analysis is therefore limited by our lack of knowledge of the demographics associated with the samples in each study.

### 2.1 Study locations

In this sub-section we summarize the data we found on each study. The studies are highly varied with differing levels of publicly available information.

#### Gangelt, Germany

This ongoing study started on March 30, 2020, and is scheduled to run through to the end of April. The study aims to determine the prevalence of antibodies in the German town of Gangelt. On April 10, 2020 the authors published preliminary results [4], stating that “around” 500 individuals had been investigated, with 14% testing positive for IgG antibodies. We assume this figure means that 70 individuals tested positive. The preprint does not mention which serology test the authors use, but the principal author is quoted in Die Zeit on April 10, 2020, stating that the study uses the IgG ELISA test produced by EUROIMMUN AG [9]. The authors do not include demographic information, but they do refer to the WHO recommendation of investigating a random sample of 100-300 households. The authors state that they investigated “approximately” 1000 individuals from “approximately” 400 households, and that the preliminary results are for “approximately” 500 individuals.

#### Geneva, Switzerland

This ongoing study started in early April 2020 in the Comté de Genève [10, 5]. The study releases results on a weekly basis. According to their study protocol, researchers use the same EUROIMMUN ELISA test for IgG antibodies as the Gangelt study. In the week of April 6 to 10, researchers found that 3.5% of the 343 participants tested positive, which we interpret as 12 individuals. In the week of April 14 to 17, they found that 5.5% of the 417 participants tested positive, which we interpret as 23 individuals.

#### Chelsea, Massachussetts

This small study is mentioned in a news piece published by Science on April 21, 2020 [2]. From the article: *Massachusetts General Hospital pathologists John Iafrate and Vivek Naranbhai quickly organized a local serology survey. Within 2 days, they collected blood samples from 200 passersby on a street corner*. Of those 200, 63 individuals tested positive. The researchers use a serological test manufactured by BioMedomics [11].

#### San Miguel County, Colorado

This study aims to test most residents from a county in Colorado, with a population of approximately 8,000. The study is conducted in partnership with the serological test manufacturer UBI Group. In a progress update published on April 21, the county announced that out of 4,757 antibody tests processed, 26 were positive, and 70 were “borderline”. The county’s web-page notes that *A borderline result on the first test indicates that the result produced a “high-signal flash” which is not enough to produce a positive result. A borderline result means that the individual may have been recently exposed to COVID-19 and may be in the early stage of producing antibodies*. In our analysis, we include the borderline cases (96 total positive tests, out of 4,757).

#### Santa Clara County, California

This study was conducted over the short period April 3, 2020 to April 4, 2020 [1]. Out of 3,330 individuals sampled, 50 tested positive suggesting a prevalence of 1.5% (exact binomial 95CI 1.11-1.97%). The authors sampled the population through targeted Facebook invitations. The resulting sample contains bias in age and socio-economic status. The authors adjusted their results by post-stratifying to match the Santa-Clara County demographics. They report an adjusted estimate for antibody prevalence of 2.81% (95CI 2.24-3.37%). This study features aCOVID-19 antibody test manufactured in China by Hangzhou Biotest Biotech Co., Ltd., and distributed in the United States by Premier Biotech, Inc.

#### Los Angeles County, California

This study was conducted between April 10, and April 11, 2020 [7, 12], involving some of the same authors from the Santa Clara study. A preprint of this study is not yet available, but the raw numbers were announced in communiques made by LA County Public Health, and University of Southern California. Out of 863 participants, 4.1% tested positive, which we interpret as 35 individuals. The researchers reported the 95% confidence bounds on the prevalence of 2.8%, and 5.6%. The sample was generated by a random draw from a marketing firm’s database, made to be representative of the County’s demographics. They used the antibody test sold by Premier Biotech, the same used for the Santa Clara study.

**Table 1:** Data from the serology studies

| Region | Test supplier | Test data |  |  |
| --- | --- | --- | --- | --- |
|  |  | Sample size | Number positive | Number negative |
| Chelsea MA | BioMedomics | 200 | 63 | 137 |
| Gangelt, Germany | EUROIMMUN | 500 | 70 | 430 |
| Geneva, Switzerland (week 1) | EUROIMMUN | 343 | 12 | 331 |
| Geneva, Switzerland (week 2) | EUROIMMUN | 417 | 23 | 440 |
| Los Angeles County | Premier Biotech | 863 | 35 | 828 |
| San Miguel County | UBI Group | 4757 | 96 | 4661 |
| Santa Clara County | Premier Biotech | 2583 | 43 | 2540 |

#### Studies not included

There are several other COVID-19 seroprevalence studies that we do not included due to a lack of data about the case counts and the serology test specifications. Given the size of these studies and the potential impact of their stated results, we feel that releasing the underlying data is of paramount importance.

- **New York**. On April 24, 2020 the State of New York announced the results of a COVID-19 antibody study conducted over the preceding week. In the sample of 3,000 individuals, the state reported that 13% tested positive. Within those, the subset of New York City residents (1,300 individuals) tested positive at 21%. We could not find any specifications about the antibody test used.
- **Netherlands**. The Netherlands’ central blood bank (Sanquin) tracks COVID-19 antibodies by sampling the blood donations they collect on a weekly basis. However, the test they use is unknown to us (one of the study leads is quoted in a Science news piece [2]: *Hans Zaaijer, a virologist at Sanquin, the Dutch national blood bank, who helped lead the study, says the team used a commercial test, which “consistently shows superior results” in validation studies, but didn’t provide more details*.. In their first set of results [8], the Dutch announced that out of 4208 samples, 135 tested positive. Of these, there were 688 (25 pos.) in the 18-30 age range, 494 (17 pos.) in the 31-40 range, 752 (26 pos.) in the 41-50 range, 1234 (38 pos.) in the 51-60 range, 1030 (29 pos.) in the 61-70 range, and 10 (0 pos.) in the 71-80 range.
- **Denmark**. Denmark is conducting a study similar to the Netherlands in which they are sampling weekly donations at their central blood bank. In an update published on April 7, 2020, the Danish Health Authority [13] indicates that among 1,000 blood donors, 2.7% had tested positive for antibodies. They also reported that the sensitivity of their test is 70% (there was no information on test specificity). This sensitivity figure is the same reported by Danish researchers for the EUROIMMUN IgG test [14]. However we could not confirm this association.

### 2.2 Test specifications

The seven studies we consider use four different test types. Each test has a different false positive rate and a different false negative rate. We include data on each test to build our models.

#### EUROIMMUN IgG ELISA test

This test is used in the Gangelt [4], and Geneva studies [10, 5]. Its performance was assessed independently by a team of Danish researchers (Lassaunière *et al*., [14]). In their assessment, 20 out of 30 confirmed positive samples tested positive, and 3 out of 82 confirmed negative samples tested positive. The authors note that “borderline data were considered negative”. These results mitigate the manufacturer’s claim that their test has >99% specificity. One reason for the lower specificity seems to be the test’s cross-reactivity with other coronaviruses, as found by Okba *et al*. [15].

#### BioMedomics COVID-19 IgM/IgG Rapid Test

The manufacturer’s own assessment [11] found that out of 397 confirmed positive samples, 352 tested positive, and out of 128 confirmed negative samples, 12 tested positive.

#### UBI SARS-CoV-2 ELISA

The manufacturer claims that *100% of the blood samples collected at day 10 or later after infection from SARS-CoV-2 from patients who tested positive to COVID-19 by other methods were also found to be positive using the UBI@ SARS-CoV-2 ELISA* [16]. In the absence of an actual figure for the number of tests, we assumed a conservative *N =* 30. In addition, UBI indicates that out of “over 900” blood samples collected before the COVID-19 outbreak, none tested positive. Again, we took a conservative approach, and assumed *N =* 900.

#### Premier Biotech COVID-19 IgG and IgM rapid test

This COVID-19 antibody test is manufactured in China by Hangzhou Biotest Biotech Co., Ltd., and distributed in the United States by Premier Biotech, Inc. It is based on a combination of IgG, and IgM antibody detection. Both the manufacturer and the researchers leading the Santa Clara County study, assessed the test performance. The manufacturer found that 75out of 75 clinically confirmed COVID-19 patients with positive IgG tested positive, and 78 out of 85 confirmed IgM-positive samples tested positive. The manufacturer also found that 369 out of 371 confirmed negative samples tested negative. The Santa Clara study authors found that in a set of 37 samples confirmed to be PCR-positive, and IgG-or IgM-positive, 25 tested positive. They also found that out of 30 confirmed negative samples, 30 tested negative.

**Table 2:** Antibody test false positive results. Population known to not have COVID-19.

| Test supplier | Test data |  |  |
| --- | --- | --- | --- |
|  | Sample size | Number positive | Number negative |
| BioMedomics | 128 | 12 | 116 |
| EUROIMMUN | 82 | 3 | 79 |
| Premier Biotech | 401 | 2 | 399 |
| UBI Group | 900 | 0 | 900 |

**Table 3:** Antibody test false negative results. Population known to have COVID-19.

| Test supplier | Test data |  |  |
| --- | --- | --- | --- |
|  | Sample size | Number positive | Number negative |
| BioMedomics | 397 | 352 | 45 |
| EUROIMMUN | 30 | 20 | 10 |
| Premier Biotech | 197 | 178 | 19 |
| UBI Group | 30 | 30 | 0 |

### 2.3 Statistical analysis and results

Our first approach uses a hierarchical Bayesian model, which we solved by Markov Chain Monte Carlo using Stan [17]. There are three observables in the model:

- *n*_obs_(*i*), the number of positive antibody tests out of *N*_obs_(*i*) individuals tested in each location i;
- *n*_fpos_(*j*), the number of positive antibody tests out of *N*_neg_(*j*) confirmed negative samples in the validation for each test brand *j* (*false positives*);
- *n*_fneg_(*j*), the number of negative antibody tests out of *N*_pos_(*j*) confirmed positive samples in the validation for each test brand *j* (*false negatives*).

From these observables, we infer, *p*_prev_(*i*), the prevalence of antibodies at each location, i. The number of positive antibody tests *n*_obs_(*i*) observed in each study are random variables that depend on *p*_prev_(*i*), but also on *p*_ipos_(*j*(*i*)) and *p*_fneg_(*j*(*i*)), which are the false positive and false negative rates of each serology test type *j*, indexed as a function of study *i*, namely

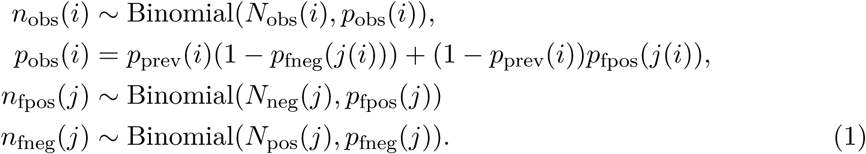

Our prior construction reads,

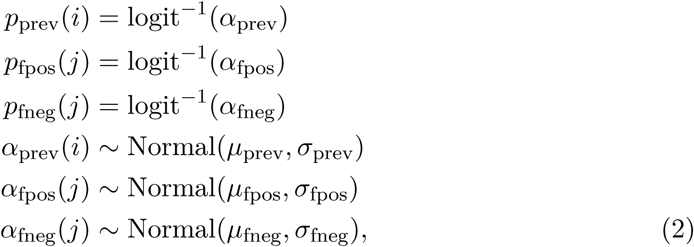

with hyper-priors of,

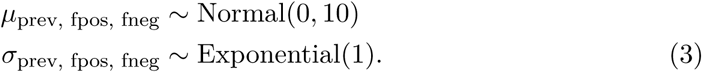

We run the model in Stan using 4 chains each with 10,000 samples, discarding the first 5,000 as part of the MCMC warm-up. The 20,000 post warm-up samples result in 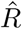 values above 0.9999 for all parameters in the model, and effective sample counts of approximately 10,000.

We plot the marginal posterior density functions for the antibody prevalence at each location in figure 1. The Santa Clara study shows a density function consistent with a high probability of a non-zero antibody prevalence, with a mean and a mode slightly greater than 1%, although we note that the posterior distribution does include zero. Los Angeles stands out—the mode of the distribution sits at 4% with a 95% credible interval that does not include zero. The German town of Gangelt was particularly hard hit by COVID-19 and the subsequent serology study suggests an antibody prevalence of 14%. Our Bayesian analysis is consistent with the study’s result, but we see that the posterior distribution has a tail that includes a prevalence below 10%.

**Figure 1:**
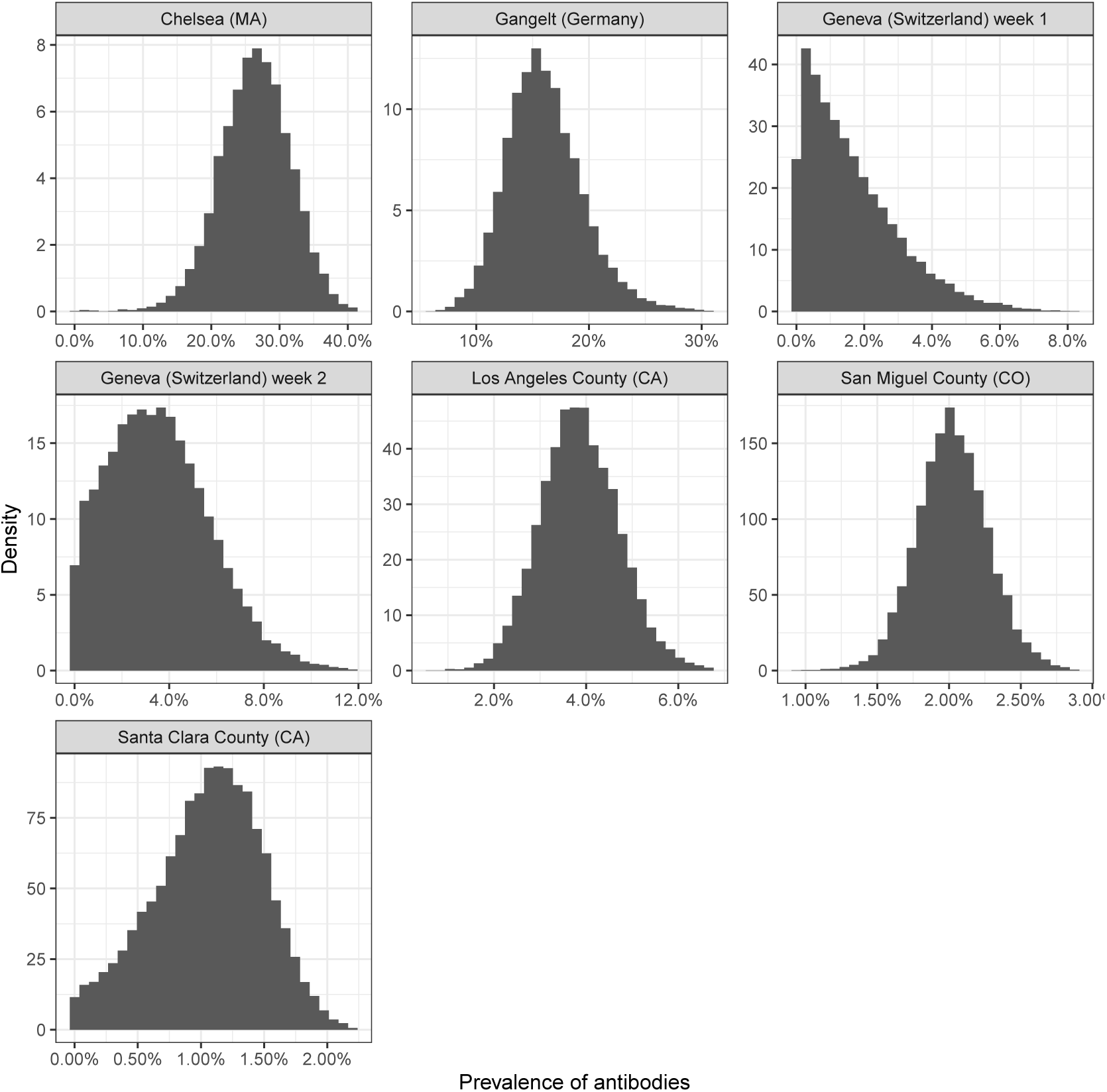
Marginal posterior distribution functions for antibody prevalence at each study location from Bayesian hierarchical model.

The contours in figure 2 show a two dimensional slice through the posterior distribution revealing the probability density in the prevalence-false positive plane. While the Santa Clara study has a false positive rate competitive with the implied prevalence, our Bayesian result clearly shows the mode located substantially greater than zero prevalence.

**Figure 2:**
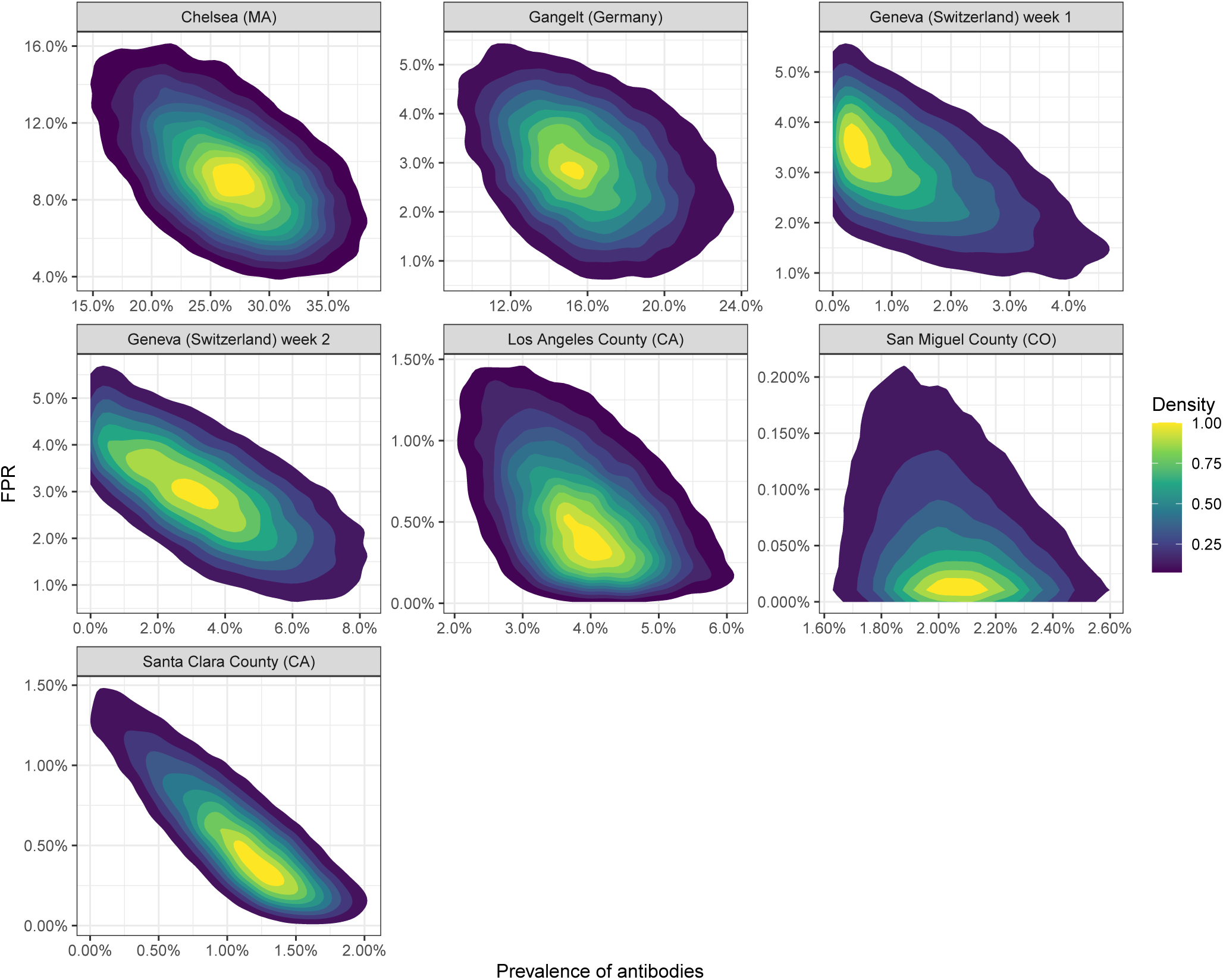
The two dimensional marginal posterior distribution functions for antibody prevalence with the false positive rate at each study location from Bayesian hierarchical model.

**Figure 3:**
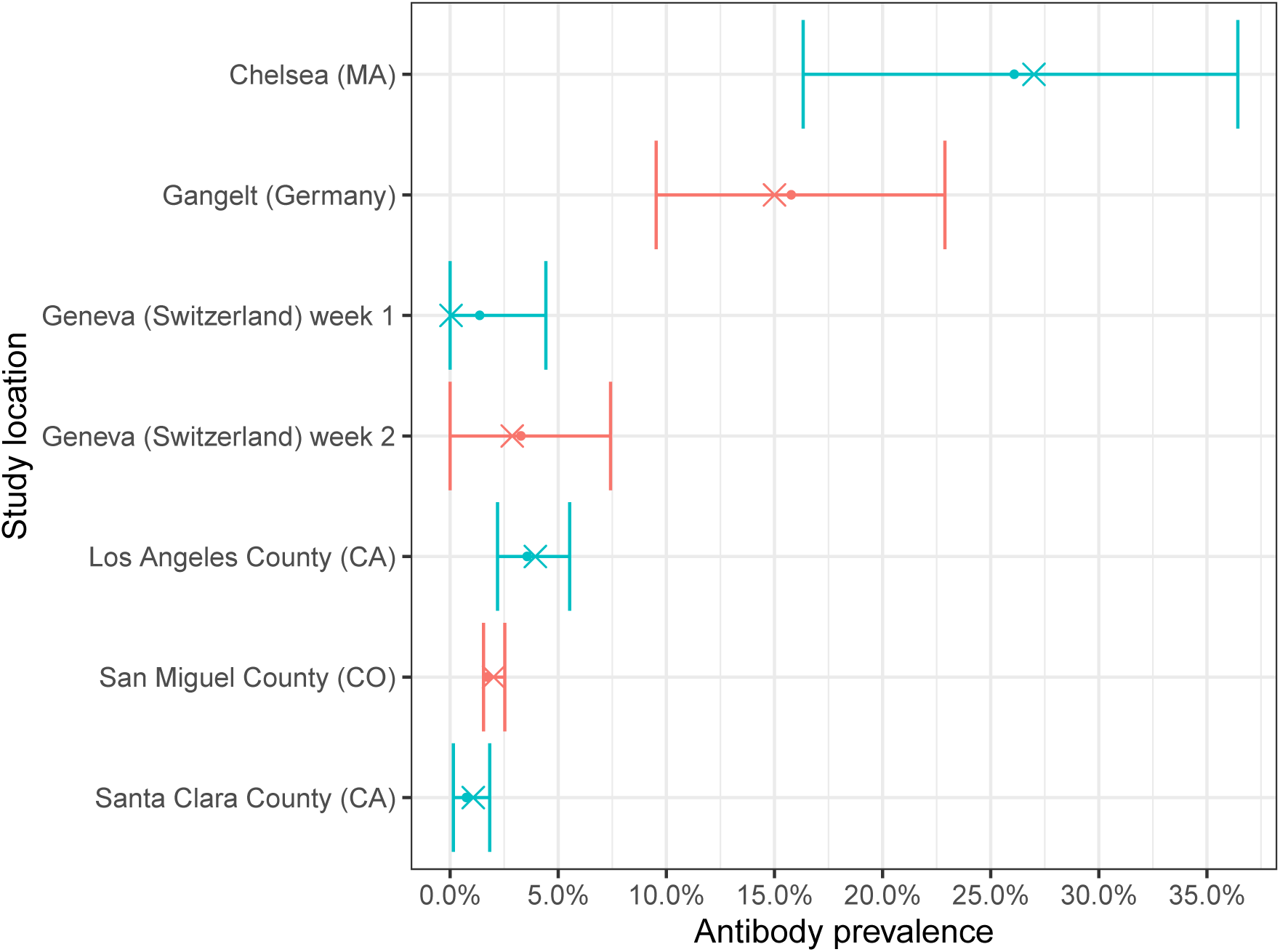
Prevalence 95% credible intervals from the Bayesian model. Circles show the mean, crosses show the mode.

Figures 1 and 2 show that Geneva has an implied prevalence close to zero. The EUROIMMUN test used in the Geneva studies has a relatively high false positive rate, creating tension in the resulting marginal posterior distributions. A significant probability mass sits at zero for both weeks of the Geneva study.

Both figures for San Miguel County (CO) show a strong antibody prevalence result. The strength comes from the low false positive rate associated with the study’s serology test (UBI). However, data on the UBI test is not fully available, and the strength of the result in part relies on our educated guess on the test’s sensitivity, as outlined in 2.2. The Bayesian analysis pulls the low false positive rate away from zero.

The Chelsea (MA) study uses a serology test (BioMedomics) with a relatively high false positive rate. However, the strength of the signal coupled to the Bayesian learning across all the studies strongly suggest a highly non-zero antibody prevalence level. The 95% credible interval around the mode excludes a prevalence level below 10%.

As a check on the the Bayesian implementation, we use a set of binomial Generalized Linear Mixed Models (GLMM) with prediction sampling. Mixed models provide estimations in situations which have have sub-population specific effects by borrowing strength from population averages (see, for example, [18]). The borrowing effect or “shrinkage” tempers those sub-populations which have relatively less data but otherwise allows the data to speak for itself. GLMM provides an optimal compromise between complete pooling and no pooling of the sub-populations in a regression analysis. In that sense GLMM is a precursor to Bayesian approaches which permit greater flexibility through priors and hyper-priors and afford more control over shrinkage effects.

Using [19, 20], we build separate GLMMs for the empirical count observation process across the study locations, for the test type false positive rates, and for the test type false negative rates. In total, we have three GLMMs. We then build Monte Carlo prediction samples [21] for the means of each GLMM and compute the implied prevalence. The GLMM specification is:

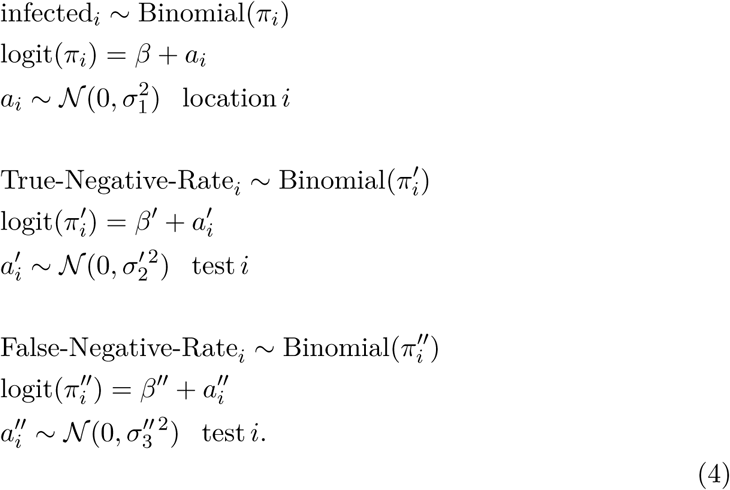

We estimate the implied prevalence distribution at each location from,

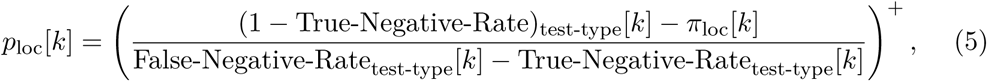

where *k* represents the *k*-th Monte Carlo sample from the GLMM prediction. The density function of *p*_loc_[*k*] is the prevalence density function at each location.

We estimate the three separate GLMMs in eq.(4) using [20] and we display the fixed and random effects with their respective the 95% confidence intervals in figures 4, 5, and 6. To build our Monte Carlo estimate of the prevalence density function in eq.(5), we sample the predictions from the models over the uncertainty in each parameter with each realization representing the mean of the model, namely

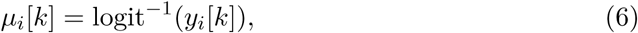

where *i* denotes the GLMM model for the *k*-th sample.

**Figure 4:**
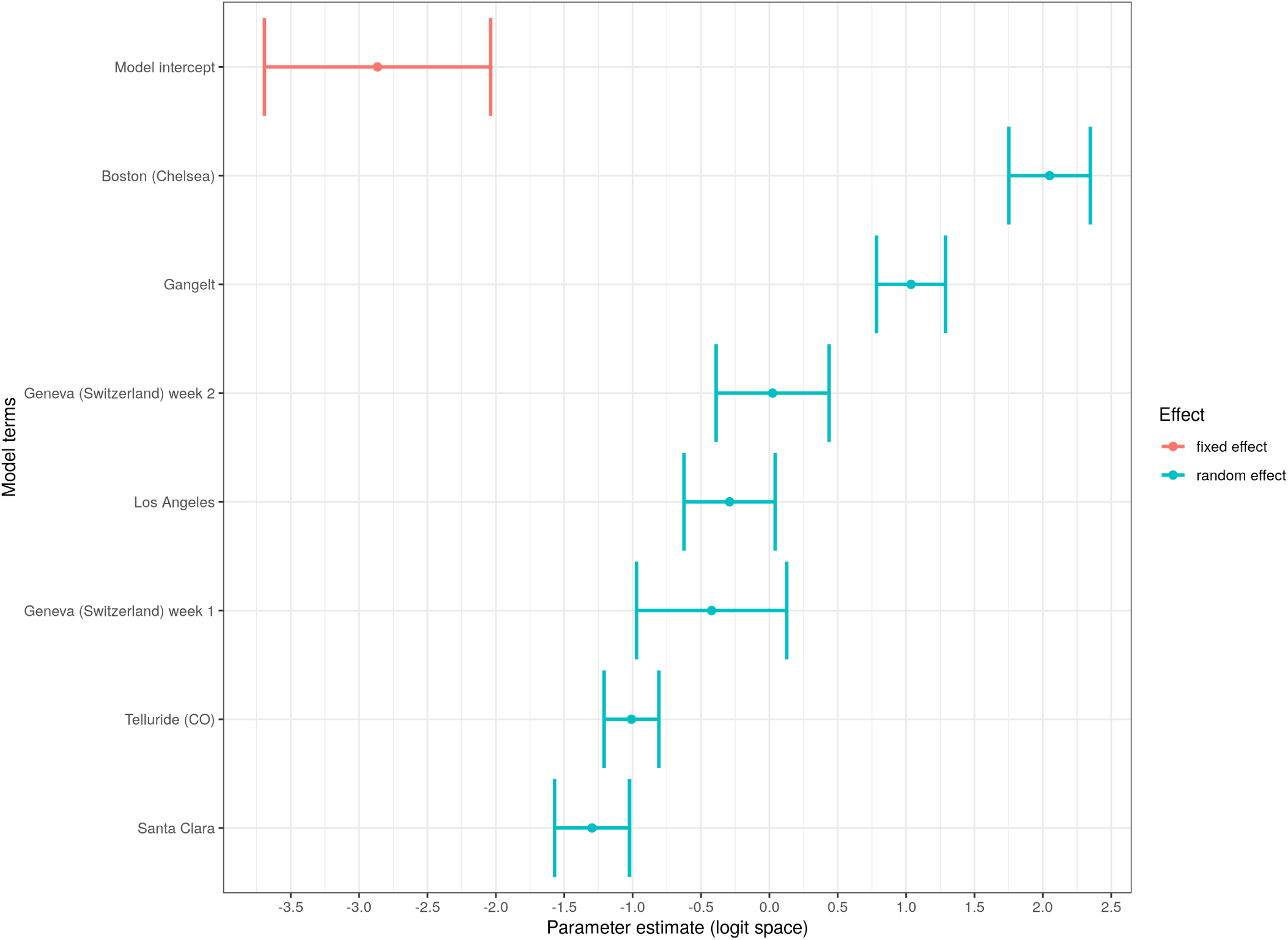
Model terms, *β* and *a_i_*, of the empirical count GLMM. The fixed and random effect terms are in logit space.

**Figure 5:**
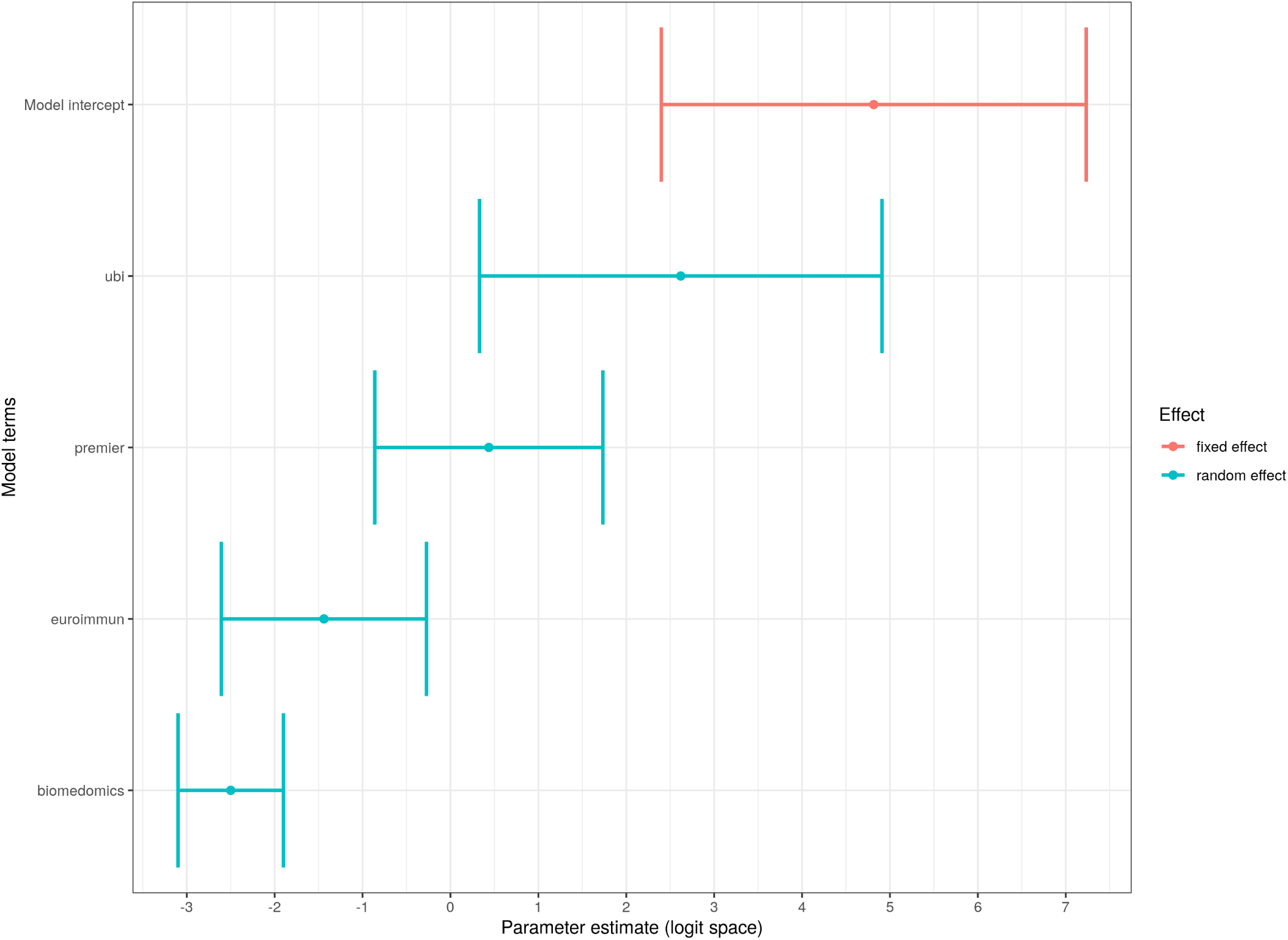
Model terms, *β*′ and 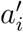, of the serology test true negative GLMM. The fixed and random effect terms are in logit space.

**Figure 6:**
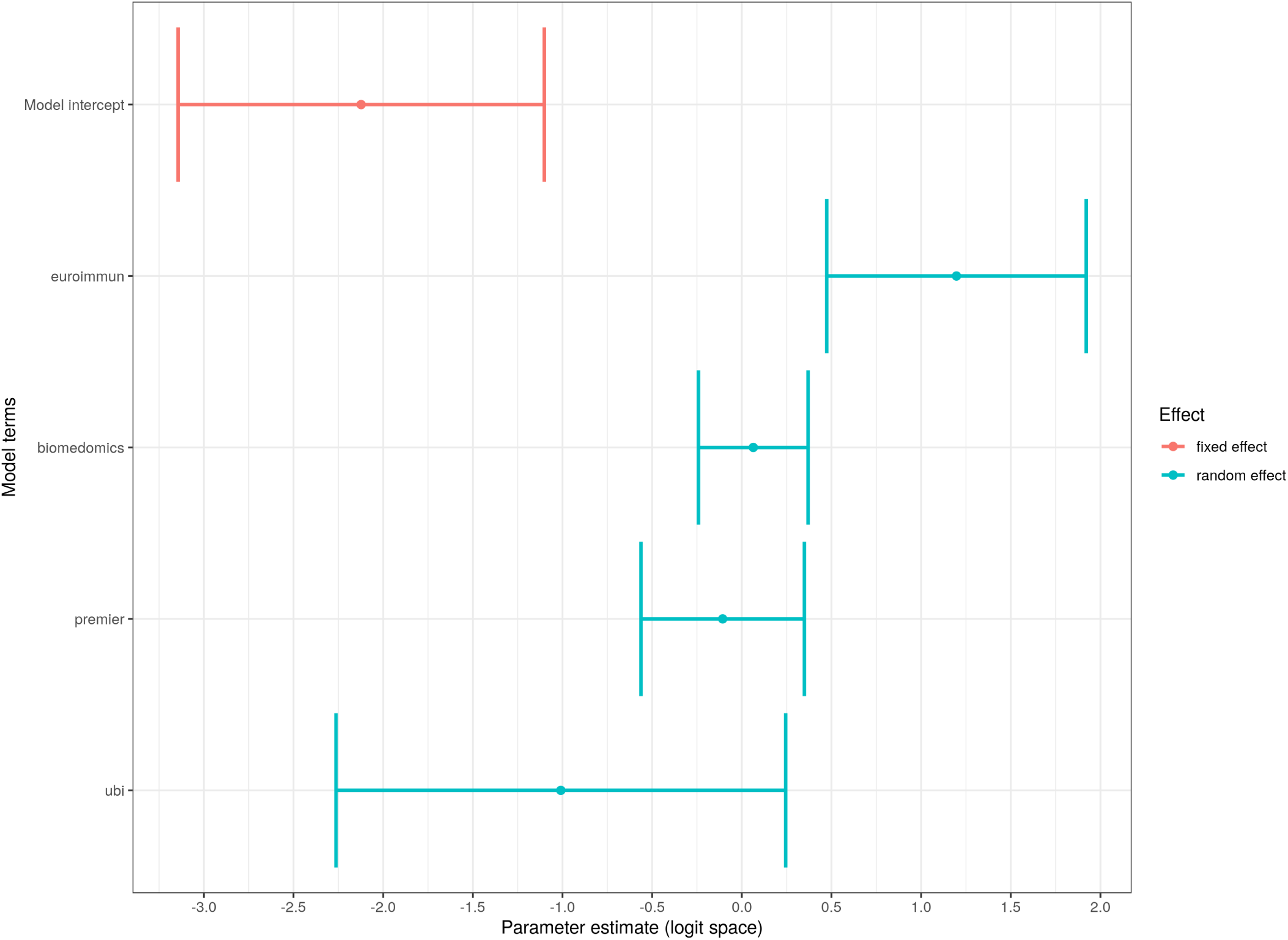
Model terms, *β*″ and 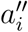, of the serology test false negative GLMM. The fixed and random effect terms are in logit space.

Figure 7 shows the resulting density functions for the antibody prevalence in each location from the GLMM prediction sampling. Notice the similarity with figure 1; both of our implementations lead to similar density functions and are in broad agreement. The Santa Clara study suggests a prevalence of 1.5% and we see in the figure that the quoted value is just beyond the mode of the distribution. We also notice that Santa Clara has a heavy tail towards zero, just as we found in the Bayesian analysis. Yet, in part by relying on “borrowing strength” from the across all the studies, we also see that the Santa Clara result clearly suggests a non-zero antibody prevalence. Again, we also see a strong effect in Los Angeles County, with a mode of 4%; the 95% confidence interval around the mean does not include zero. In figure 8 we show the 95% confidence intervals with the mean for all locations computed from the GLMM sampling.

**Figure 7:**
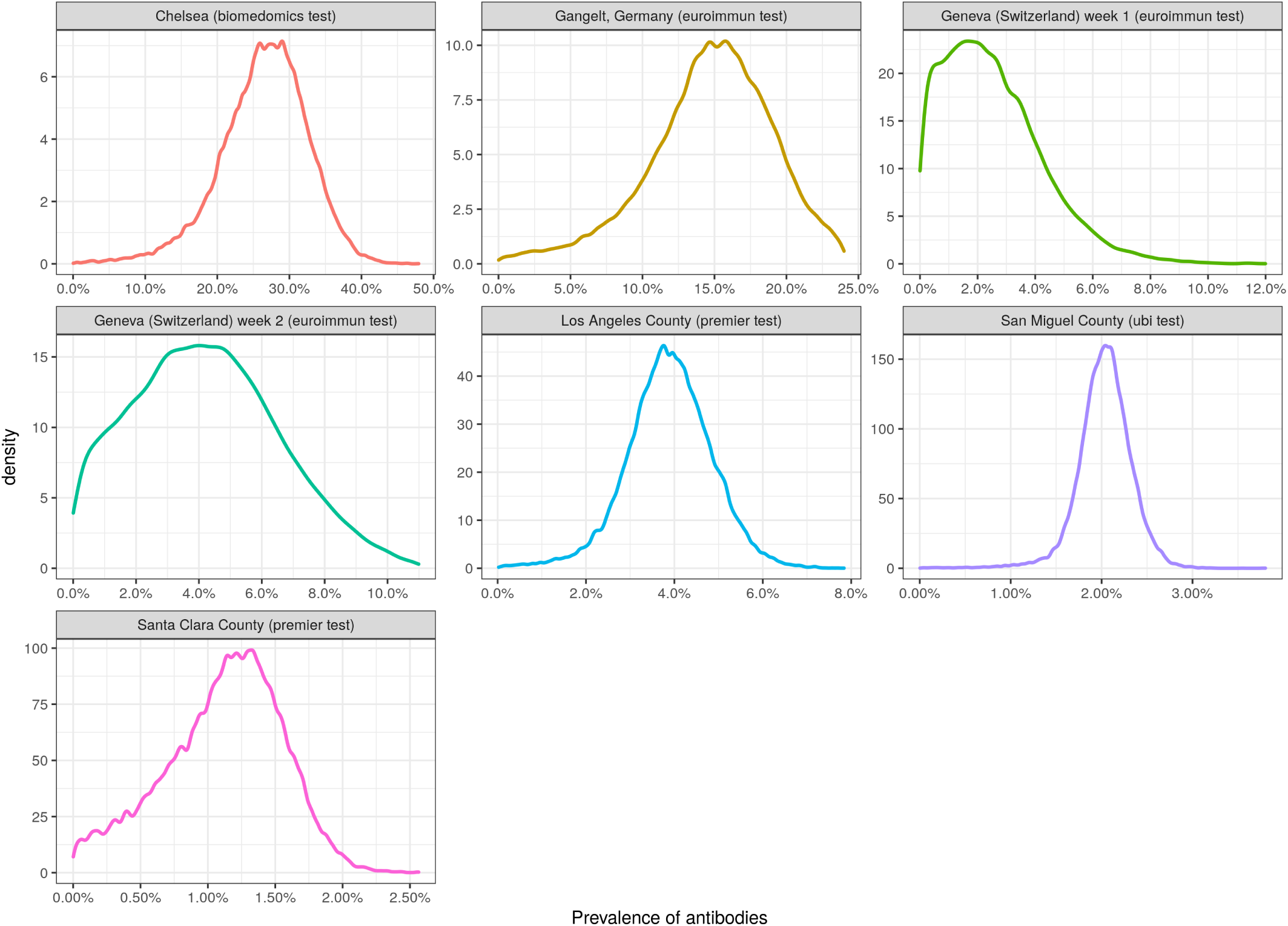
Density functions for antibody prevalence at each study location from the GLMM construction.

**Figure 8:**
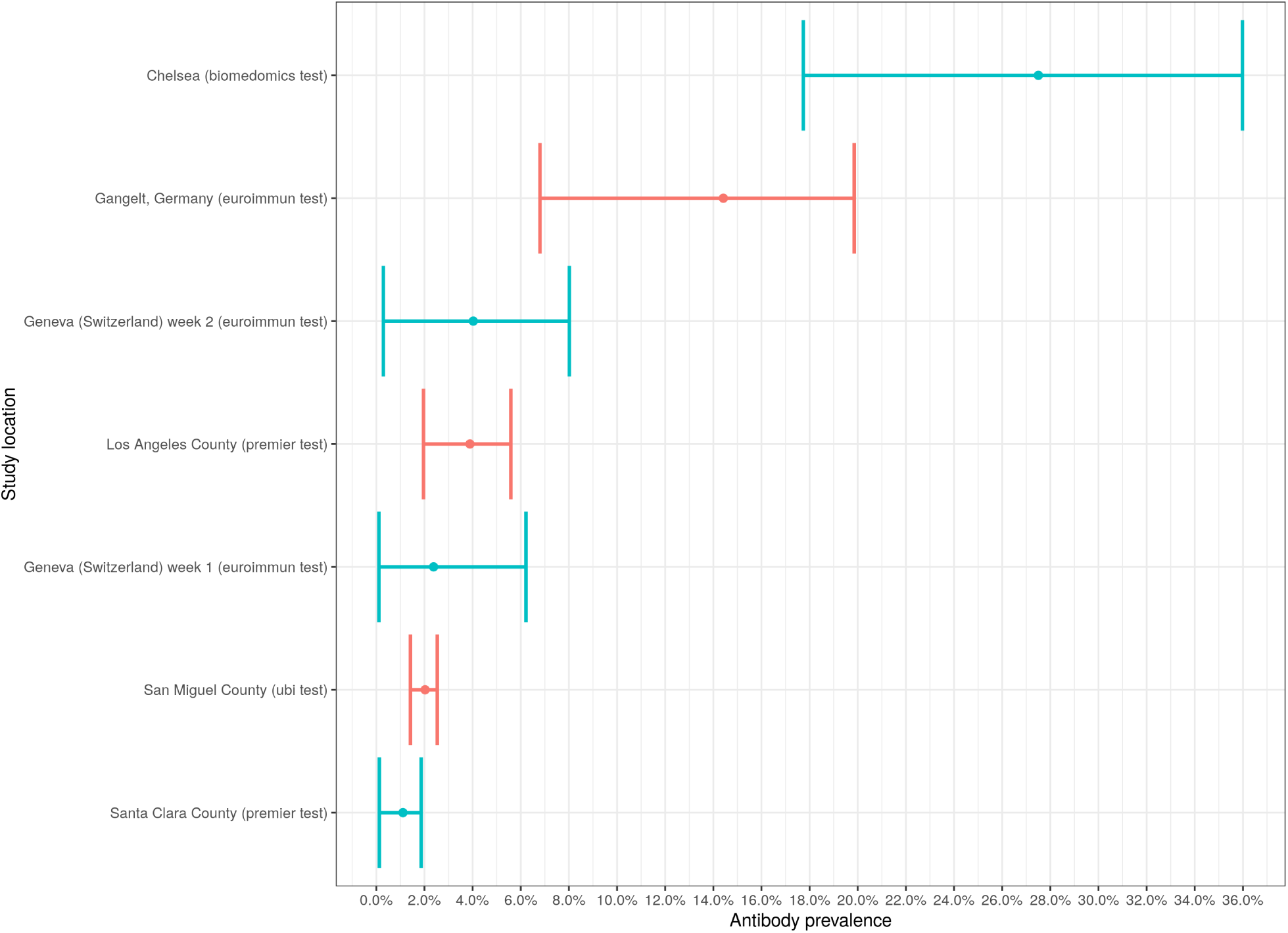
Prevalence 95% confidence intervals for each study location from the GLMM construction. Mean indicated.

In figure 9 we show a contour density map of the prevalence with the false positive rate. We can clearly see the tension between the false positive rate and prevalence but we also see a clear signal associated with each region. Again, notice the similarity to figure 2. In particular, note that San Miguel County (CO) again shows a strong prevalence result, but that the GLMM shrinks the false positive rate away from zero.

**Figure 9:**
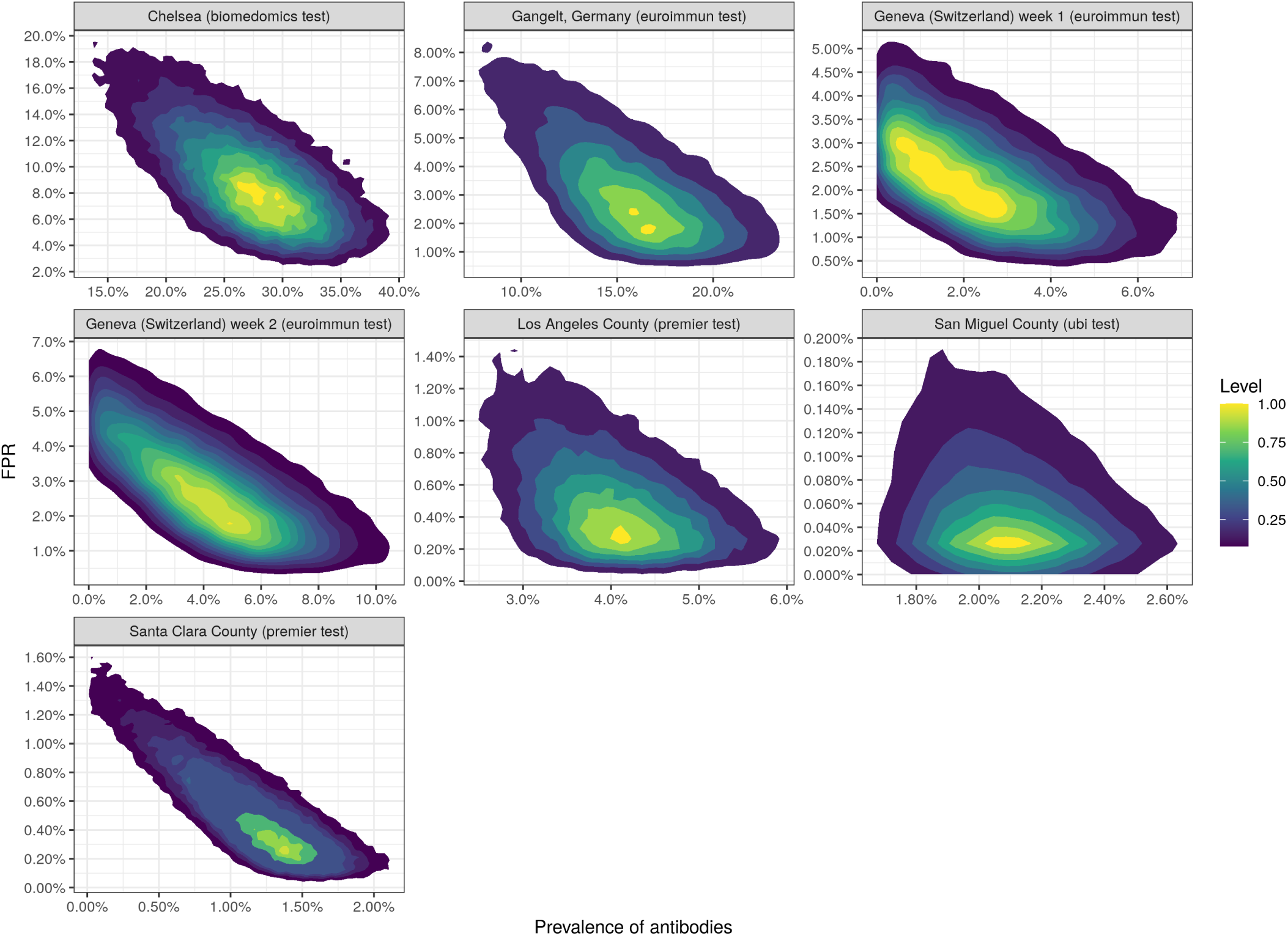
Two dimensional density functions for antibody prevalence with the false positive rate at each study location from the GLMM construction.

The GLMM is in some sense “semi-Bayesian”—we have a hierarchical model with a random effect on the intercept term in logit space which we sample over, but we do not have a formal set of priors with hyper-parameters. While the GLMM provides a basis for learning across the data, our full Bayesian specification with Markov Chain Monte Carlo provides a more complete result. However, by implementing the GLMM we see that our results are robust to different approaches and model specifications. Our results are not sensitive to the specifics of the prior and hyper-prior constructions in our Bayesian model.

Both methods demonstrate that there is significant evidence for non-trivial antibody prevalence in the populations associated with these studies. While the individual statistical power of each study is not high, using Bayesian techniques and GLMM constructions on the data from all the studies reveal a definite signal that we should take seriously.

## 3 Discussion

Seroprevalence studies are an important tool in combating COVID-19 since public policies are dependent on how far the disease has already penetrated into the general population. Not only does serology testing help us understand the overall infection fatality rate of the disease, but testing also helps us design targeted strategies such as contact tracing. Furthermore, serology studies can provide insight into the dynamics of the disease propagation.

While we do not correct for possible population sample bias or other demographic issues, our analysis points to an important signal: the seroprevalence studies to date show that a significant fraction of the populations examined have antibodies against SARS-CoV-2 in their bloodstream. The exact prevalence levels are highly region dependent. As more serology studies appear, they will sharpen our understanding of antibody prevalence in the general population.

The quality of antibody prevalence estimates depend on sample size and on the specificity/sensitivity of the antibody test. High test specificity increases statistical power. Thus, there is a natural trade-off between investing in tests of high quality, the human resources required to generate large sample sizes, and the speed at which society needs results. The societal and economic consequences between Type I and Type II errors are not equal.

## Data Availability

All data and data sources are included in the manuscript.

## Notes

### Competing Interest Statement

The authors have declared no competing interest.

### Funding Statement

No external funding.

